# Sodium-glucose cotransporter-2 inhibitors and the risk of lung cancer among patients with type 2 diabetes: a retrospective cohort study

**DOI:** 10.1101/2022.08.09.22278566

**Authors:** Samantha B. Shapiro, Hui Yin, Oriana H.Y. Yu, Laurent Azoulay

**Affiliations:** Department of Epidemiology, Biostatistics, and Occupational Health, McGill University, Montreal, Canada; Center for Clinical Epidemiology, Lady Davis Institute, Jewish General Hospital, Montreal, Canada; Division of Endocrinology, Jewish General Hospital, Montreal, Canada; Gerald Bronfman Department of Oncology, McGill University, Montreal, Canada

## Abstract

**Objective:** To determine whether the use of sodium-glucose cotransporter 2 (SGLT-2) inhibitors, compared to dipeptidyl peptidase 4 (DPP-4) inhibitors, is associated with a decreased risk of incident lung cancers among patients with type 2 diabetes.

**Methods:** We assembled a new-user, active comparator cohort of SGLT-2 inhibitor and DPP-4 inhibitor users using the United Kingdom Clinical Practice Research Datalink. We fit Cox proportional hazards models with propensity score fine stratification weighting to estimate hazard ratios (HRs) with 95% confidence intervals (CI) for incident lung cancers.

**Results:** Crude lung cancer incidence rates were 0.92 and 1.37 per 1,000 person-years among 47,517 SGLT-2 inhibitor and 129,807 DPP-4 inhibitor users, respectively. No reduced risk of lung cancer was observed among SGLT-2 inhibitor users after weighting (HR 1.08, 95% CI 0.78–1.48). Conclusions: In this large cohort study, the use of SGLT-2 inhibitors was not associated with a decreased risk of lung cancer.

## INTRODUCTION

Laboratory studies have demonstrated that premalignant lesions and adenocarcinomas of the lung have increased expression of SGLT-2 [1, 2]. SGLT-2 inhibitors may therefore have a beneficial role in the prevention and treatment of lung cancer, potentially by affecting cell cycle arrest and apoptosis [3]. Animal models have shown that administering canagliflozin to mice with premalignant lesions of the lungs resulted in delayed tumour onset, a lower tumour burden, and prolonged survival, while administering canagliflozin to mice with lung adenocarcinoma patient-derived xenografts resulted in reduced tumour volume [1]. However, no studies to date have investigated the association between the use of SGLT-2 inhibitors and lung cancer risk in humans. We conducted a retrospective cohort study to assess whether SGLT-2 inhibitor use, when compared to dipeptidyl peptidase-4 (DPP-4) use, is associated with a decreased risk of lung cancer in individuals with type 2 diabetes.

## METHODS

To investigate this relationship, we sourced data from the United Kingdom (UK) Clinical Practice Research Datalink (CPRD) [4]. We assembled a cohort of patients newly prescribed an SGLT-2 inhibitor (canagliflozin, empagliflozin, dapagliflozin, ertugliflozin) or a DPP-4 inhibitor (alogliptin, linagliptin, saxagliptin, sitagliptin, vildagliptin) between 1 January 2013 (the year when SGLT-2 inhibitors became available in the UK) and 31 December 2019. Cohort entry was the date of the first-ever prescription for an SGLT-2 inhibitor or DPP-4 inhibitor during the study period. We excluded patients less than 18 years of age, with less than one year of medical history, with a history of any cancer other than non-melanoma skin cancer, or with less than 6 months of follow-up.

Patients were followed starting six months after cohort entry to account for latency of the effect of SGLT-2 inhibitor use on lung cancer. Patients were censored upon the first occurrence of an incident lung cancer diagnosis, death from any cause, end of registration with the CPRD, or end of the study period (30 June 2020).

Our models considered a broad range of potential confounders measured at or before cohort entry, including age, sex, calendar year, body mass index, smoking status, hemoglobin A1c, alcohol-related disorders, duration of diabetes, previous use of antidiabetic drugs, proxy variables for diabetes severity such as macrovascular and microvascular complications, history of lung diseases, common comorbidities, medication use, and markers for health-seeking behaviour such as cancer screening and vaccination. A missing variable category was used for missing covariate data as missing data was unlikely to be differential between exposure groups.

We generated propensity scores for all participants using multivariable logistic regression to predict the probability of using SGLT-2 inhibitors compared to DPP-4 inhibitors conditional on all covariates listed in Table 1. We used propensity score fine stratification weighting to control for confounding and generated 50 strata of patients based on the propensity score distribution of the SGLT-2 inhibitor users. We used descriptive statistics to assess balance between groups using standardised differences and considered differences of less than 0.10 to be acceptable.

**Table 1:** Baseline characteristics of SGLT-2 inhibitor and DPP-4 inhibitor exposure groups after weighting.

| Characteristics | SGLT-2 inhibitors | DPP-4 inhibitors | Standardized difference |
| --- | --- | --- | --- |
| Total | 47,517 | 129,807 |  |
| Age, years, mean (SD) | 55.9 (10.8) | 55.9 (11.1) | 0.00 |
| Male, n (%) | 28,321 (59.6) | 76,759 (59.1) | 0.01 |
| Year of cohort entry, n (%) |  |  |  |
| 2013 | 843 (1.8) | 2,292 (1.8) | 0.00 |
| 2014 | 3,702 (7.8) | 9,433 (7.3) | 0.02 |
| 2015 | 6,398 (13.5) | 16,428 (12.7) | 0.02 |
| 2016 | 7,347 (15.5) | 19,698 (15.2) | 0.01 |
| 2017 | 8,416 (17.7) | 23,242 (17.9) | 0.01 |
| 2018 | 9,337 (19.6) | 25,924 (20.0) | 0.01 |
| 2019 | 11,474 (24.1) | 32,791 (25.3) | 0.03 |
| Body mass index, mean (SD) |  |  |  |
| BMI <30 kg/m<sup>2</sup>, n (%) | 11,341 (23.9) | 31,492 (24.3) | 0.01 |
| BMI ≥30.0 kg/m<sup>2</sup>, n (%) | 35,665 (75.1) | 96,875 (74.6) | 0.01 |
| Unknown | 511 (1.1) | 1,440 (1.1) | 0.00 |
| Smoking status, n (%) |  |  |  |
| Ever | 36,369 (76.5) | 99,370 (76.6) | 0.00 |
| Never | 11,134 (23.4) | 30,397 (23.4) | 0.00 |
| Unknown | 14 (0.0) | 40 (0.0) | 0.00 |
| Hemoglobin A1c, n (%) |  |  |  |
| ≤53 mmol/mol (7.0%) | 2,565 (5.4) | 7,179 (5.5) | 0.01 |
| 54–64 mmol/mol (7.1%–8.0%) | 9,962 (21.0) | 26,981 (20.8) | 0.00 |
| >64 mmol/mol (8.0%) | 34,803 (73.2) | 95,126 (73.3) | 0.00 |
| Unknown | 187 (0.4) | 520 (0.4) | 0.00 |
| Alcohol-related disorders, n (%) | 3,999 (8.4) | 11,117 (8.6) | 0.01 |
| Duration of diabetes in years, mean (SD) | 8.3 (6.3) | 8.2 (6.2) | 0.02 |
| Previous use of antidiabetic drugs, n (%) |  |  |  |
| Metformin | 44,307 (93.2) | 120,964 (93.2) | 0.00 |
| Sulfonylureas | 17,101 (36.0) | 46,816 (36.1) | 0.00 |
| Thiazolidinedione | 3,291 (6.9) | 9,254 (7.1) | 0.01 |
| Meglitinides | 151 (0.3) | 493 (0.4) | 0.01 |
| Alpha-glucosidase inhibitors | 69 (0.1) | 171 (0.1) | 0.00 |
| GLP-1 RAs | 6,184 (13.0) | 15,956 (12.3) | 0.02 |
| Insulin | 8,934 (18.8) | 24,506 (18.9) | 0.00 |
| Macrovascular complications, n (%) |  |  |  |
| Peripheral vascular disease | 3,111 (6.5) | 8,693 (6.7) | 0.01 |
| Stroke | 1,391 (2.9) | 3,858 (3.0) | 0.00 |
| Myocardial infarction | 2,844 (6.0) | 7,627 (5.9) | 0.00 |
| Microvascular complications, n (%) |  |  |  |
| Renal disease | 2,715 (5.7) | 8,311 (6.4) | 0.03 |
| Retinopathy | 16,264 (34.2) | 43,182 (33.3) | 0.02 |
| Neuropathy | 8,618 (18.1) | 23,263 (17.9) | 0.01 |
| History of lung disease, n (%) |  |  |  |
| COPD | 4,001 (8.4) | 10,893 (8.4) | 0.00 |
| Interstitial lung disease | 62 (0.1) | 169 (0.1) | 0.00 |
| Comorbidities, n (%) |  |  |  |
| Heart failure | 1,022 (2.2) | 3,015 (2.3) | 0.01 |
| Non-alcoholic fatty liver disease | 2,238 (4.7) | 6,162 (4.7) | 0.00 |
| Sleep apnea | 3,250 (6.8) | 8,686 (6.7) | 0.01 |
| Osteoarthritis | 9,172 (19.3) | 25,295 (19.5) | 0.00 |
| Depression | 19,097 (40.2) | 52,114 (40.1) | 0.00 |
| Medication use, n (%) |  |  |  |
| ACE inhibitors | 21,474 (45.2) | 57,899 (44.6) | 0.01 |
| Angiotensin receptor blockers | 7,458 (15.7) | 20,339 (15.7) | 0.00 |
| Beta blockers | 9,543 (20.1) | 26,107 (20.1) | 0.00 |
| Calcium channel blockers | 13,335 (28.1) | 36,283 (28.0) | 0.00 |
| Diuretics | 7,280 (15.3) | 20,158 (15.5) | 0.01 |
| Antiarrhythmic agents | 1,482 (3.1) | 3,983 (3.1) | 0.00 |
| Antiplatelet agents | 11,205 (23.6) | 30,241 (23.3) | 0.01 |
| Statins | 35,185 (74.0) | 94,910 (73.1) | 0.02 |
| Proton pump inhibitors | 17,165 (36.1) | 46,805 (36.1) | 0.00 |
| NSAIDs | 10,087 (21.2) | 27,633 (21.3) | 0.00 |
| Opioids | 14,948 (31.5) | 41,427 (31.9) | 0.01 |
| Health-seeking behaviours, n (%) |  |  |  |
| Faecal occult blood testing or colonoscopy | 6,574 (13.8) | 17,977 (13.8) | 0.00 |
| Mammography | 3,155 (6.6) | 8,686 (6.7) | 0.00 |
| Prostate-specific antigen testing | 2,640 (5.6) | 7,275 (5.6) | 0.00 |
| Influenza vaccination | 700 (1.5) | 1,887 (1.5) | 0.00 |
| Pneumococcal vaccination | 2,772 (5.8) | 7,459 (5.7) | 0.00 |

*Abbreviations: SD, standard deviation; DPP-4, dipeptidyl peptidase 4; SGLT, sodium-glucose cotransporter; GLP-1 RA, glucagon-like peptide-1 receptor agonist; COPD, chronic obstructive pulmonary disease; NSAID, non-steroidal anti-inflammatory drug*

We calculated incidence rates of lung cancer and 95% confidence intervals (95% CIs) based on the Poisson distribution in both exposure groups over the follow-up period. We fit weighted Cox proportional hazards models to estimate the hazard ratio (HR) and 95% CI of lung cancer incidence in SGLT-2 inhibitor users compared to DPP-4 inhibitor users. We additionally assessed whether there is a duration-response relationship between SGLT-2 inhibitor use and lung cancer incidence, whether the relationship between SGLT-2 inhibitor use and lung cancer incidence varies within the individual drugs in the class, and whether smoking acts as an effect modifier for the relationship between SGLT-2 inhibitor use and lung cancer. Finally, we conducted four sensitivity analyses: 1) varying the six-month lag period to twelve-month and eighteen-month periods, 2) conducting an as-treated analysis such that patients were censored upon drug discontinuation or prescription of a drug from the opposite exposure group, 3) using GLP-1 receptor agonist users as the comparator group rather than DPP-4 inhibitor users, and 4) using inverse probability of censoring weighting to account for potential differences in mortality and censoring between the two exposure groups.

The study was approved by the CPRD Independent Scientific Advisory Committee (protocol 22_001710).

## RESULTS

We followed 47,517 SGLT-2 inhibitor users for a median of 2.00 years and 129,807 DPP-4 inhibitor users for a median of 2.84 years (Figure 1). The cohort was well balanced with respect to covariates (Table 1). The crude incidence rates of lung cancer were 91.88 and 136.98 per 100,000 person-years among SGLT-2 inhibitor users and DPP-4 inhibitor users, respectively (Table 2). After weighting, no decreased risk of lung cancer was observed among SGLT-2 inhibitor users (HR 1.08, 95% CI 0.78–1.48).

**Figure 1:**
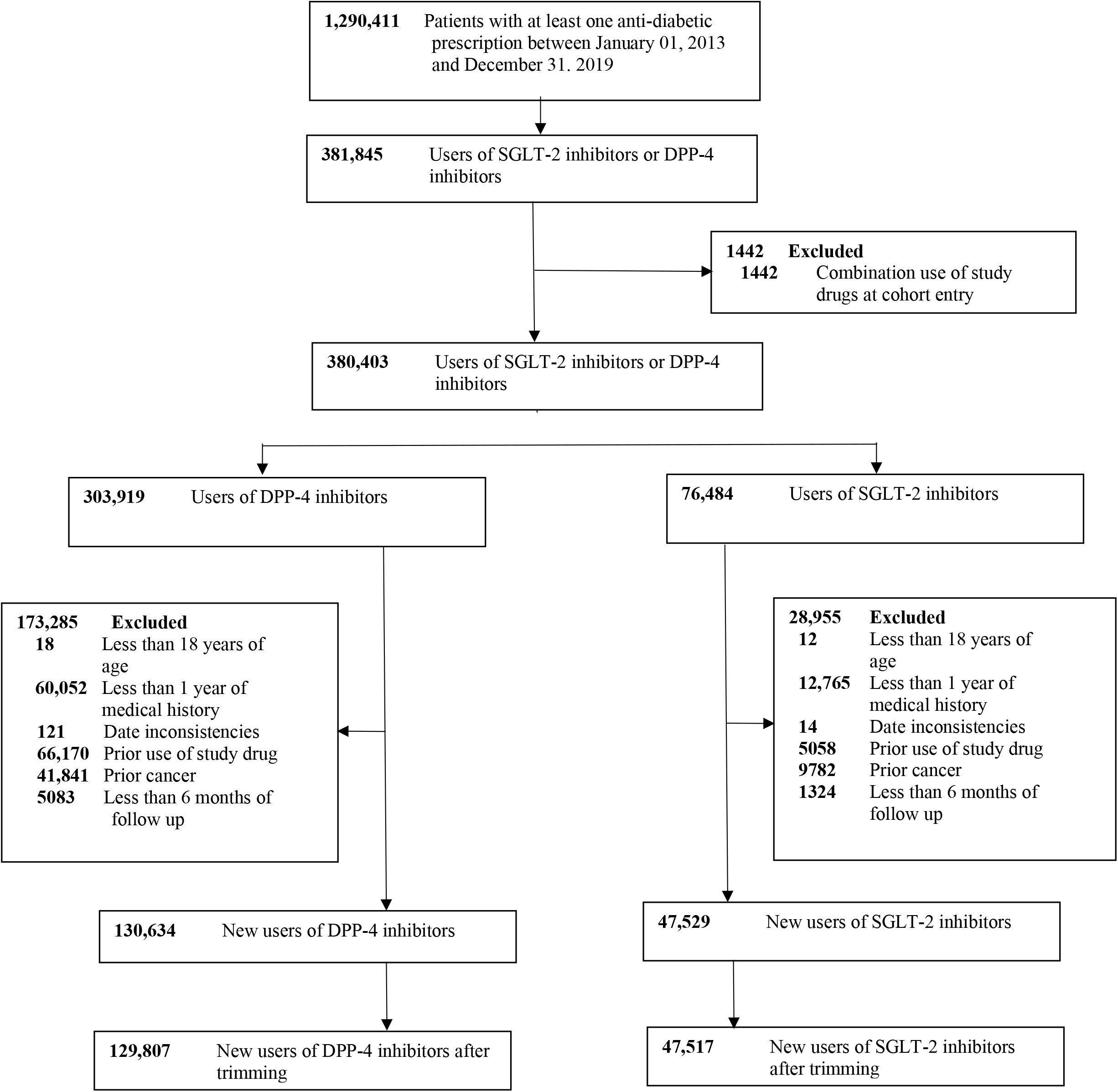
Study flow chart.

**Table 2:** Hazard ratios for lung cancer comparing SGLT-2 inhibitors with DPP-4 inhibitors.

| Analysis | Events | Person-years | Crude IR<sup>a</sup> | Weighted IR (95% CI)<sup>a</sup> | Crude HR (95% CI) | Weighted HR (95% CI) |
| --- | --- | --- | --- | --- | --- | --- |
| Primary analysis<sup>b</sup> |  |  |  |  |  |  |
| DPP-4 inhibitors | 542 | 395,680 | 136.98 | 85.18 (74.89–96.50) | 1.00 [Reference] | 1.00 [Reference] |
| SGLT-2 inhibitors | 100 | 108,843 | 91.88 | 91.88 (74.75–111.74) | 0.69 (0.55–0.85) | 1.08 (0.78–1.48) |
| Cumulative use<sup>b,c</sup> |  |  |  |  |  |  |
| <6 months | 24 | 27,926 | 85.94 | 85.94 (55.06–127.87) | 0.65 (0.43–0.99) | 0.99 (0.59–1.67) |
| 6–12 months | 20 | 24,623 | 81.23 | 81.23 (49.62–125.45) | 0.61 (0.39–0.96) | 0.98 (0.58–1.64) |
| >12 months | 56 | 56,295 | 99.48 | 99.48 (75.14–129.18) | 0.73 (0.56–0.97) | 1.17 (0.81–1.69) |
| Type of SGLT-2 inhibitor<sup>d,e</sup> |  |  |  |  |  |  |
| Canagliflozin | 20 | 18,265 | 109.50 | 109.50 (66.89–169.11) | 0.84 (0.53–1.31) | 1.22 (0.71–2.10) |
| Dapagliflozin | 52 | 61,982 | 83.90 | 83.90 (62.66–110.02) | 0.63 (0.47–0.83) | 0.93 (0.62–1.38) |
| Empagliflozin | 28 | 28,554 | 98.06 | 98.06 (65.16–141.72) | 0.77 (0.52–1.14) | 1.30 (0.81–2.09) |
| Ertugliflozin<sup>f</sup> | 0 | 10 | - | - | - | - |
| Effect measure modification<sup>d,g</sup> |  |  |  |  |  |  |
| Non-smokers | 3 | 24,340 | 12.33 | 12.33 (2.54–36.02) | 0.54 (0.16–1.84) | 0.66 (0.14–3.14) |
| Ever-smokers | 97 | 84,490 | 114.81 | 114.81 (93.10–140.05) | 0.70 (0.57–0.88) | 1.10 (0.80–1.51) |
| Sensitivity analyses |  |  |  |  |  |  |
| Varying lag period<sup>d,h</sup> |  |  |  |  |  |  |
| 12-month lag | 87 | 87,167 | 99.81 | 99.81 (79.94–123.11) | 0.73 (0.58–0.92) | 1.14 (0.81–1.60) |
| 18-month lag | 61 | 68,768 | 88.70 | 88.70 (67.85–113.94) | 0.66 (0.50–0.87) | 1.10 (0.66–1.58) |
| As-treated<sup>b,i</sup> | 55 | 70,945 | 77.52 | 77.52 (58.40–100.91) | 0.57 (0.43–0.76) | 0.98 (0.70–1.39) |
| Alternate comparator<sup>b,j</sup> | 191 | 183,374 | 104.16 | 104.16 (89.91–120.02) | 1.06 (0.82–1.36) | 1.02 (0.64–1.63) |
| IPCW<sup>k,l</sup> | 100 | 122,354 | 81.73 | 78.79 (69.89–88.52) | 0.66 (0.53–0.82) | 0.99 (0.71–1.37) |

Abbreviations: IR, incidence rate; HR, hazard ratio; CI, confidence interval; DPP-4, dipeptidyl peptidase 4; SGLT, sodium-glucose cotransporter, GLP-1 RA, glucagon-like peptide-1 receptor agonist; IPCW, inverse probability of censoring weighting
<sup>a</sup> Per 100,000 person-years
<sup>b</sup> Models were weighted using propensity score fine stratification
<sup>c</sup> DPP-4 inhibitor users (reference) generated 542 events over 395,680 person-years
<sup>d</sup> Models were weighted using propensity score fine stratification with separate propensity score analyses conducted for each comparison
<sup>e</sup> DPP-4 inhibitor users (reference) generated 382 events over 295,060 person-years
<sup>f</sup> PS model was not created for the comparison between ertugliflozin and DPP-4 inhibitors
<sup>g</sup> DPP-4 inhibitor users (reference) generated 21 events over 81,918 person-years among non-smokers and 521 events over 313,665 person-years among ever-smokers
<sup>h</sup> DPP-4 inhibitor users (reference) generated 466 events over 334,396 person-years with a twelve-month lag period and 386 events over 278,907 person-years with an eighteen-month lag period
<sup>i</sup> DPP-4 inhibitor users (reference) generated 338 events over 245,433 person-years
<sup>j</sup> GLP-1 RA users (reference) generated 91 events over 88,474 person-years and were followed for a median of 3.07 years
<sup>k</sup> Models were weighted using propensity score fine stratification and inverse probability of censoring weighting
<sup>l</sup> DPP-4 inhibitor users (reference) generated 542 events over 433,550 person-years

There was little evidence of a duration-response relationship. For cumulative durations of less than six months, six to twelve months, and greater than twelve months of SGLT-2 inhibitor use, weighted hazard ratios were 0.99 (95% CI 0.59–1.67), 0.98 (95% CI 0.58–1.64), and 1.17 (95% CI 0.81–1.69), respectively. The relationship between SGLT-2 inhibitor use and lung cancer did not vary by SGLT-2 inhibitor molecule (HR 1.22 (95% CI 0.71–2.10) for canagliflozin, HR 0.93 (95% CI 0.62–1.38) for dapagliflozin, HR 1.30 (95% CI 0.81–2.09) for empagliflozin). Effect modification by smoking was not evident (HR 0.66 (95% CI 0.14–3.14) among non-smokers, HR 1.10 (95% CI 0.80–1.51) among ever-smokers).

Our estimates remained consistent when varying the lag period from six months to twelve months (HR 1.14, 95% CI 0.81–1.60) and eighteen months (HR 1.10, 95% CI 0.66–1.58). There was little effect of censoring patients upon drug discontinuation or prescription of a drug from the opposite exposure group (HR 0.98, 95% CI 0.70–1.39). Estimates remained similar when comparing SGLT-2 inhibitor users to GLP-1 receptor agonist users instead of DPP-4 inhibitor users (HR 1.02, 95% CI 0.64–1.63). Finally, our estimates were similar when using inverse probability of censoring weighting (HR 0.99, 95% CI 0.71–1.37).

## CONCLUSIONS

This large cohort study was the first to assess the relationship between SGLT-2 inhibitor use and the risk of lung cancer in humans. The results of this study do not suggest that the use of SGLT-2 inhibitors, compared to DPP-4 inhibitors, is associated with a decrease in the short-term risk of lung cancer. Further research on this association with a longer follow-up period may be warranted.

## Supporting information

STROBE reporting checklist

## Data Availability

No data are available. This study is based on data from the Clinical Practice Research Datalink obtained under license from the UK Medicines and Healthcare products Regulatory Agency. The data are provided by patients and collected by the UK National Health Service as part of their care and support. The interpretation and conclusions contained in this study are those of the author/s alone. Because electronic health records are classified as "sensitive data" by the UK Data Protection Act, information governance restrictions (to protect patient confidentiality) prevent data sharing via public deposition. Data are available with approval through the individual constituent entities controlling access to the data. Specifically, the primary care data can be requested via application to the Clinical Practice Research Datalink (https://www.cprd.com).

## Author contributions

S.B.S. wrote the first draft of the manuscript, and all authors edited, reviewed, and approved the final version of the manuscript. All authors conceived and designed the study, analyzed and interpreted the data, approved the final version of the manuscript, and are accountable for its accuracy. S.B.S., H.Y., and L.A. performed the statistical analyses. L.A. acquired the data and supervised the study. L.A. is the guarantor of this work and, as such, had full access to all the data in the study and takes responsibility for the integrity of the data and the accuracy of the data analysis.

## REFERENCES

[1] Scafoglio CR, Villegas B, Abdelhady G, et al. (2018) Sodium-glucose transporter 2 is a diagnostic and therapeutic target for early-stage lung adenocarcinoma. Sci Transl Med 10(467): eaat5933. 10.1126/scitranslmed.aat5933

[2] Ishikawa N, Oguri T, Isobe T, Fujitaka K, Kohno N (2001) SGLT gene expression in primary lung cancers and their metastatic lesions. Jpn J Cancer Res 92(8): 874–879. 10.1111/j.1349-7006.2001.tb01175.x

[3] Zhou J, Zhu J, Yu S-J, et al. (2020) Sodium-glucose co-transporter-2 (SGLT-2) inhibition reduces glucose uptake to induce breast cancer cell growth arrest through AMPK/mTOR pathway. Biomed Pharmacother 132: 110821. https://doi.org/10.1016/j.biopha.2020.110821

[4] Herrett E, Gallagher AM, Bhaskaran K, et al. (2015) Data Resource Profile: Clinical Practice Research Datalink (CPRD). Int J Epidemiol 44(3): 827–836. 10.1093/ije/dyv098

